# Exploring the Impact of a Medical Device Recall on Individuals with Obstructive Sleep Apnea and Healthcare Providers: A Qualitative Study

**DOI:** 10.64898/2026.03.25.26349320

**Authors:** Sachin R. Pendharkar, Kenneth G. Blades, Bashour Yazji, Najib T. Ayas, Robert L. Owens, Marta Kaminska, Constance Mackenzie, Andrea S. Gershon, Diana C. Ratycz, Vadym Lischenko, Mark E. Fenton, Kerry A. McBrien, Marcus Povitz, Tetyana Kendzerska

## Abstract

**Purpose:** To understand how the Philips PAP device recall affected patient experiences, clinical practice, and health system responses.

**Methods:** From November 2022 to August 2023, we interviewed individuals with OSA, physicians, respiratory therapists and health system leaders. We also received emailed responses from Health Canada. Interviews explored participants’ experiences with the recall announcement and communication, their own responses and perceptions of actions taken by others, the overall impact of the recall and suggestions for improving future recall processes. Interviews were analyzed using an inductive thematic approach.

**Results:** We interviewed 47 participants (16 individuals with OSA, 10 physicians, 17 public or private respiratory therapists, five health system leaders). Themes were organized into four domains: recall communication, execution, participant experiences, and the policy and regulatory context. Participants were confused due to inadequate information from Philips throughout the process. The burden of notifying patients and tracing devices mostly fell to healthcare providers and vendors, while replacement efforts were disorganized and frustrating. Individuals with OSA experienced emotional distress over therapy decisions and difficulties navigating the recall. Healthcare providers described moral distress from being unable to support patients adequately, and vendors faced additional logistical and financial strain. While regulatory authorities reported that Philips followed standard procedures, participants expressed a loss of trust in both the manufacturer and oversight systems.

**Conclusions:** Interviews revealed that poor communication and execution of the Philips recall caused confusion, frustration and significant emotional and financial burden. Collaborative, context-specific strategies are required to improve future recalls.

## INTRODUCTION

Obstructive sleep apnea (OSA) is a highly prevalent, treatable chronic disease with significant medical consequences including poorer quality of life, elevated cardiovascular risk, and disproportionate use of health system resources compared to the general population.[1–9] Societal impacts include motor vehicle and workplace accidents, and lost productivity.[10–12] The most widely used treatment for OSA is positive airway pressure (PAP) therapy, which mitigates many complications of untreated OSA and is cost effective.[8,9,13–17]

On June 14, 2021, Philips Respironics, one of the largest PAP device manufacturers globally, issued a voluntary recall notification (United States) and Field Safety Notice (International) for nearly all models of PAP devices and ventilators manufactured before April 26, 2021, impacting 15 million devices worldwide.[18] The recall was due to degradation of polyester-based polyurethane (PE-PUR) sound abatement foam, which was suspected of causing irritant and toxic effects through: (a) inhalation or ingestion of particulate matter in air tubing, and (b) inhalation of volatile organic compounds. Potential exposure risks included irritation of the skin, eyes, and respiratory tract, hypersensitivity and inflammatory reactions, headaches, and toxic or carcinogenic effects.[19]

The United States Food and Drug Administration (FDA) classified the recall as the most serious and urgent type (Class I). Health Canada issued a recall notice on June 23, 2021, indicating a similar level of risk (Type I).[20] Philips Respironics initially advised patients using recalled PAP devices for non-life threatening indications to discontinue use,[21] but subsequently revised the recommendation to encourage patients to seek advice from their physicians about device use.[22] Health Canada also advised that treatment not be discontinued prior to discussion with a healthcare provider.[23] By January 31, 2024, the FDA had received over 116,000 adverse event reports related to recalled PAP devices and 561 reports of death suspected or confirmed to be related to PE-PUR foam breakdown.[24] Simultaneously, several medical societies released statements highlighting the potential harms of untreated OSA if PAP therapy was discontinued.[21,22]

Despite these serious recall-related risks, the experiences and perspectives of patients and providers are only partially understood. In a prior survey of sleep clinicians, respondents reported that the recall negatively impacted their patients’ health and wellbeing and reduced patients’ trust in medicine.[25] Clinicians also reported uncertainty, confusion, concern for patients, and increased workload related to the recall.[25] In a recent survey of 632 Canadians with self-reported OSA, over 40% of respondents reduced or discontinued use of recalled PAP devices, 46% experienced emotional or mental health impacts and 15% described financial burden from the recall. Moreover, 31-54% reported loss of trust in manufacturers, suppliers and providers.[26] These results highlight the wide-ranging effects of the recall on patients and healthcare providers; however, the survey methodology limited the participants’ ability to describe experiences in greater depth and restricted opportunities to capture the richness of perspectives or identify novel themes beyond those anticipated by the researchers. The objective of this study was to conduct an in-depth qualitative exploration of how the recall, and its impacts, were experienced by individuals with OSA, healthcare providers, PAP vendors and health system leaders.

## METHODS

### Study Design

This was a qualitative descriptive study using semi-structured interviews with individuals with OSA, physicians, respiratory therapists and health system leaders in Canada. Interviews were conducted between November 2022 and August 2023. Ethics approval was obtained from the University of Calgary Conjoint Health Research Ethics Board (REB22-0803), the Ottawa Hospital Research Institute (Ethics ID: 20220590-01T), and Clinical Trials Ontario (Ethics ID: CTO-4064).

### Study Setting

In Canada, provinces and territories are responsible for healthcare funding and delivery, leading to variable models of OSA care.[27,28] Physician assessments are covered through provincial public health insurance. Individuals suspected to have OSA may be referred for ambulatory or laboratory-based sleep diagnostic testing, which are provided in most provinces through a mix of public and private facilities. PAP devices are available by prescription from community-based vendors mostly staffed by respiratory therapists (RTs) or sleep technicians. Funding for PAP therapy varies by province, ranging from full public funding to completely private funding which requires patients to bear the cost of therapy (either out-of-pocket or through private insurance). In all provinces, public funding is available for individuals with low-income, significant disability, or complex respiratory sleep disorders requiring advanced non-invasive ventilation. Medical devices such as PAP machines are regulated federally, but decisions about manufacturer may be based on provincial government or vendor contracts depending on the funding model.

### Study Participants

We recruited participants from the main groups affected by or involved in the recall process: individuals with OSA, physicians, respiratory therapists (RTs), and health system leaders. Individuals with OSA were adults reporting a physician diagnosis of OSA who were currently using or had previously used a PAP device. Physicians were sleep specialists from academic and community settings who managed individuals with OSA in their clinical practice. Respiratory therapists practiced in public (e.g., publicly funded sleep laboratory) and private settings (e.g., respiratory equipment vendor). Furthermore, we recruited RTs who worked directly with patients in clinical settings (“clinical”) and those serving in administrative roles such as managers, directors or vendor owner/operators (“administrative”). Health system leaders included officials from government programs and other non-government healthcare organizations and provider associations.

Participants were recruited through email newsletters and social media posts distributed by healthcare organizations and patient associations, posters and pamphlets placed in sleep laboratories and respiratory equipment vendor clinics, and university research participation websites (see online supplement). A research associate contacted individuals who responded to these advertisements to obtain informed consent. Participants were also asked to provide our study contact information to colleagues or others whose participation they thought would be relevant. We have previously used these recruitment methods to obtain a diversity of perspectives.[29] We requested an interview from Health Canada, the federal agency responsible for medical device regulation. They declined our request but provided written responses to questions we submitted and engaged in subsequent follow-up communications. No other policy agencies replied to our emailed interview requests.

### Data Collection

Interviews were conducted by a trained investigator experienced with qualitative research methods and knowledgeable about the recall (KGB). Interview guides were developed by our multidisciplinary team, comprising people with lived experience of OSA, specialist and primary care physicians, and health services researchers. Interview guide topics included how participants learned about the recall, their actions during the recall, personal/professional impacts of the recall, their assessment of the decisions and actions of other groups involved in the recall, and suggestions for improvement of recall processes in the future. These topics were chosen to cover the scope of the recall process and to prompt detailed description of experiences. The interviewer also explored unanticipated topics when they arose during interviews. Interviews were offered in English and French, although all participants chose to be interviewed in English. Each participant was offered a gift card as a token of appreciation. All interviews were recorded and transcribed verbatim.

### Data Analysis

Two investigators (KGB and BY) used an inductive thematic approach to analyze the interview data in consultation with the co-principal investigators (TK and SRP). Analysis began with low-level open coding of concrete data elements such as actions, events, gaps, barriers, and dates and timelines. Each interview was coded comprehensively by both analysts, first in parallel and subsequently independently once the inductive codebook was determined to be saturated. The analysts then engaged in a second inductive and iterative process to identify high-level themes and sub-themes representing recurring patterns across the dataset. The analysts conferred regularly to discuss their engagement with the data, refine their approach, and made analytic decisions collaboratively and with continual reference to the primary data. We used NVivo version 14 (Lumivero LLC, Denver, CO, USA) to store and organize interview transcripts and to facilitate coding.

## RESULTS

We interviewed 47 participants from eight Canadian provinces or territories: 10 sleep physicians, 17 RTs (public sector (n=4), vendor (n=12) or both (n=1)) working in clinical and/or administrative roles, five health system leaders, and 16 individuals with OSA (all of whom had used recalled devices). Individuals with OSA were mostly male (69%) and 94% had moderate or severe OSA (See Table 1).

**Table 1.**
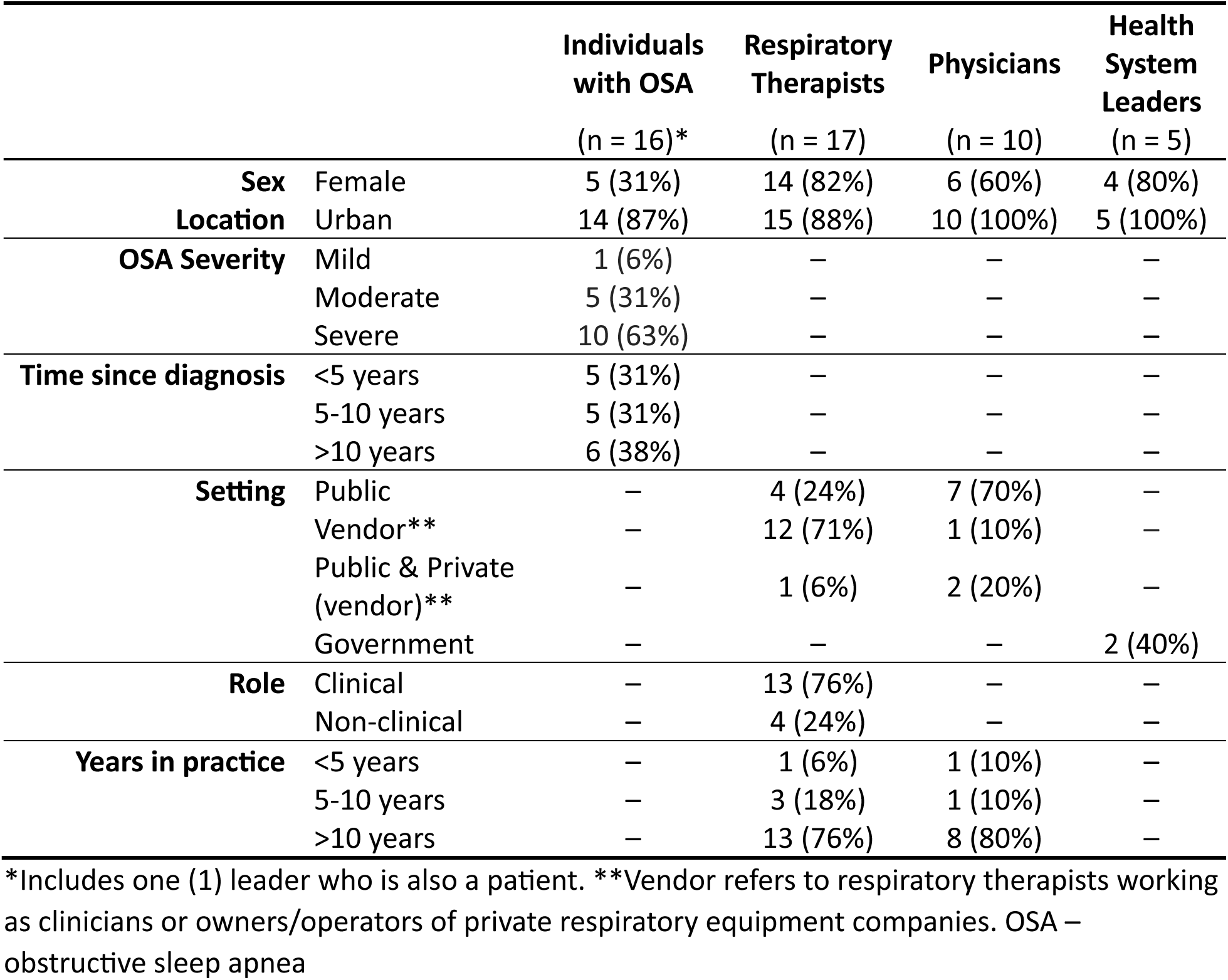
Participant characteristics.

Emerging themes were grouped into four domains: (1) communication about the recall, (2) execution of the recall, (3) experience of the recall, and (4) policy and regulatory context. Tables 2-5 demonstrate themes and representative quotes for each domain.

**Table 2.** “Communication about the recall” domain.

| **Sub-domain** | **Illustrative Quotes** |
| --- | --- |
| *Announcement and early information* | It was very vague, it was very much, “oh, go talk to your doctor, go talk to your doctor.” It wasn’t like we really had anything to really go off of. There was an internal memo with our company about it. And it was basically just, “talk to your doctors. Go on the physician’s advice.” (Interview 15, Vendor RT (Clinical))<br>The level of risk was unclear [...] Depending who you talk to, they’re like, “The risk is relatively low to relatively high.” It’s high because it’s chronic. It’s low because no-one is aware that any acute side effects have happened on patients. And so it was too wide to know what the risk was. (Interview 46, Health System Leader) |
| *Role of Philips in communication* | I never would have imagined such a mess of confusion. No information, down to the [Philips] reps who are supposed to be communicating with us: them throwing their hands in the air not knowing what happened, that was – it was disgraceful. Truly. (Interview 2, Vendor RT (Administrative))<br>I went online, looked up the model number, and it’s a heavy U.S.-based website. They [Philips] mentioned Canada, Health Canada hadn’t approved the recall yet, they were still working with the government. That’s over a year ago, nothing further from them. (Interview 3, Individual with OSA)<br>By Christmas of last year I felt really dumb because every single time a patient was calling me, I’d be telling them something different about the recall and they’re like, “But you told me this.” I’m like, “Mm-hmm, like yes, yes, I did. [...] They’ve [Philips] actually changed the story like three times since then and so take this information now with a grain of salt because it might be true and it might change again” [...] (Interview 4, Vendor RT (Clinical))<br>Manufacturers must notify consignees (anyone who received, purchased or used the affected product) in a timely manner concerning the recall and the actions to be undertaken associated with the affected devices. Manufacturers’ Quality Agreements with other parties in the supply chain, including wholesalers, distributors or storage sites, should clearly define the responsibilities for each party in the recall strategy. [...] The recalling firm must also follow up with anyone who does not respond to the initial recall notification. For type I recalls, there must be 100% contact. (Health Canada, from written statement provided to researchers) |
| *Alternative information sources to fill gaps* | It was anxiety-provoking not having enough information, but eventually the expert medical committees made recommendations [...] (Interview 7, Sleep Physician)<br>I started a WhatsApp group for the [provincial] sleep physicians. [...] By the time July/June 2021 rolled around we’re probably a good year into this established group, and so I put it [the recall notice] into the WhatsApp chat and the rest blew up. So people who weren’t informed may have become informed through that messaging system [...] (Interview 41, Sleep Physician) |

**Table 3.** “Recall execution” domain.

| **Sub-domain** | **Illustrative Quotes** |
| --- | --- |
| *Notification and traceability* | We did phone calls with a lot of people who we know don’t text, so we didn’t have their information or their consent to contact them that way. But like I say, as the weeks go by, we only have four staff members, we’re not a big company that has lots of resources in order to, you know, develop this. So, yeah, we phoned people for about three weeks, and then eventually just had to make the call and like, “We’re not really getting anywhere with this. We’re not calling nearly enough people and everybody you call –” Each phone call was like a good 10 minutes of them panicking going, “Well what am I supposed to do? Like, where do I go from here? What’s the problem? Do you have a loaner machine? You sold me this machine, what can you do for me?” And it’s like, “Nothing. Like we’d love to, we want to, we literally have no stock to do it.” [...] So we started off doing phone calls, and a month later we still had like 500 people to call. So, we said we can’t keep doing this. So, we ended up just doing a mail out. (Interview 4, Vendor RT (Clinical))<br>[...] if you’ve ever done a mass email campaign, 5% opening of your emails is a great number. So, we really had – I mean I was pretty thrilled with 60% of actually receiving and 40% opening the emails [...] (Interview 2, Vendor RT (Administrative)) |
| *Device replacement* | [...] from my perspective as a patient, I submitted my device on their website, and then it was crickets. [...] It felt like it went into the void and I heard nothing from them until [...] two weeks ago I received a call from the vendor saying that I would be receiving a – they told me I would be receiving a new machine, but they replaced the motor. (Interview 11, Individual with OSA/Health System Leader)<br>[...] a lot of patients are not technology savvy to say, to follow the steps on the web link and register their machines. So service, customer service, plain customer service has been very weak. (Interview 9, Sleep Physician)<br>In Canada, they’re [Philips] telling them [patients] – and I mean they’re still getting told, “Your machine will be mailed to you.” No, it’s not going to be mailed to you in Canada. [...] that wasn’t even being done properly. (Interview 23, Vendor RT (Administrative)) |

**Table 4.** “Experience of the recall” domain.

| **Sub-domain** | **Illustrative Quotes** |
| --- | --- |
| *Individuals with OSA* | I think emotionally and psychologically, it's been terrible. Because yes, I'm still up in the air and wondering what's happening. So, yeah, definitely that does affect me. (Interview 32, Individual with OSA)<br>You get later at night with this machine on wondering if it's leaving little bits of foam into my lungs but, you know, how can you sleep with that going on? (Interview 47, Individual with OSA)<br>[...] everything gets worse because of my lack of sleep, you know. [...] whereas the CPAP machine would fix all that. But just, you know, it's very, you know, you go with door number one or door number two and, you know, take the consequences. (Interview 47, Individual with OSA) |
| *Physicians & Public Sector RTs* | I remember having day sheets, you know, looking at my schedule and having, you know, a clinic of 35 patients and 15 of them, the topic of the follow-up was 'Philips recall.' (Interview 5, Sleep Physician)<br>It was definitely difficult, frustrating. We like to help our patients. We're there for our patients and obviously I got to deal with the calls where they were very frustrated and where the clerks couldn't handle it. So it was one bad call after the next [...] (Interview 40, Public Sector RT (Administrative))<br>[...] it's a sad moment, right, because it's something that I cannot do anything about it. I only can provide with empathy and see where we can move on from now on, but the harm is done. And it's, this is how it affects me. (Interview 35, Sleep Physician) |
| *Vendors (Private Sector RTs)* | We had a lot of people just get super angry at us. Blame us. Write nasty reviews about us online because we didn't have answers for them. (Interview 15, Vendor RT (Clinical))<br>I mean our secretary left. Quit. I don't blame her. I mean, every day she was in tears, people coming in. It was very, very stressful and it's still very stressful. (Interview 23, Vendor RT (Administrative))<br>It's a small company, we only have two offices. We did have three but one closed down during the pandemic, just some difficulties on top of difficulties, thanks to what we're talking about today. (Interview 4, Vendor RT (Clinical)) |

**Table 5.** “Policy and regulatory context” domain.

| **Sub-domain** | **Illustrative Quotes** |
| --- | --- |
| *Trust* | We have had a couple of patients refuse to take the new Respiroics unit, they just do not want anything to do with Respiroics anymore, that they’ve broken trust with. I mean, some have left us for that reason, too. We’ve lost clients, because we carried the machine that was affected [...] (Interview 17, Vendor RT (Clinical))<br>I lost a lot of trust in the machine – in the – this company, for sure. But like I said earlier, I won’t buy a Philips product ever again. I won’t buy a razor, I won’t buy anything with the name [...] (Interview 33, Individual with OSA) |
| *Transparency & Regulatory Effectiveness* | [...] it seems to me that they [Philips] must have had data for some time and there was just not being upfront for the things they can do, the things they know and the things that they can do. There’s no transparency. (Interview 9, Sleep Physician)<br>The lack of transparency in detail about the recall, the scope of the recall, and the plan of action by the manufacturer in the timeline they will remediate the device, I think that’s what’s their [patients’] main concern. (Interview 31, Vendor RT (Clinical))<br>Philips provided their recall strategy to Health Canada which includes their expected time frames for key activities in a recall, including identifying affected clients, communicating to customers, correcting the device and follow up. [...]<br>Health Canada assesses the accuracy and relevancy of information received from companies and ensures that the information is complete, as required by sections 64-65 of the Medical Devices Regulations. The expectation of the type and relevancy of information shared with Health Canada is detailed in Section 8.2.1 of the Guide to Recall of Medical Devices (GUI-0054).<br>Phillips [sic] has provided all the information as required in section 64 of the MDR. They expect to submit information required in section 65 of the MDR when the recall is complete.<br>(Health Canada, from written statement provided to researchers)<br>I think the regulations themselves could be improved. [...] the regulations are, you have to notify people, and you have to encourage them to stop taking those devices, essentially, [...] There doesn’t seem to be any middle ground to compel a company, whether it be Philips, or Ford, or any other company where there’s a recall, to do right by the consumer or the user. Other than from the safety perspective of, “stop |
|  | using this device, there's a recall." Now a lot is left to the discretion of [...] the company that is doing the recall. (Interview 14, Health System Leader)<br>[...] the Canadian government required that it had to go through a distributor. So if we wouldn't have had to go and program every machine and call every patient and do all that – but we're doing all the work. We are doing all the work for Philips, in my opinion. [...] And we're taking all the heat. Whereas in the US and those places, all they have to say is, "Call Respironics, they're the ones dealing with it. Call Respironics, they're dealing with it." (Interview 23, Vendor RT (Administrative)) |

### Domain 1: Communication about the recall

#### Recall Announcement and Early Information

Participants described the time following the recall announcement as one of uncertainty and confusion. While some vendors suspected something unusual was happening in the weeks before the manufacturer’s official announcement, most participants were taken by surprise. Vendors were rapidly overloaded with phone calls and visits from patients. Staff found it challenging to handle this influx of calls, having themselves just learned about the recall and lacking opportunity to prepare. Most of the confusion was due to limited information presented in the manufacturer’s announcement.

> The announcement told us very little. Indicated that it potentially affected all Trilogies, which is one of the units involved, and A40 devices. They didn’t give us a serial number range. They didn’t tell us what the complications could be in terms of symptoms. They just basically said that there was a mass recall. (Interview 15, Vendor RT (Clinical))

#### Philips’ Role in Communication

Following the recall announcement, participants unanimously considered Philips’ communication to be poor. There was a shared perception that the communications strategy was driven by legal rather than patient priorities. Participants with OSA reported that the Philips recall website was confusing and did not adequately explain what had happened or what they should do. Some contacted Philips directly and reported long wait times for Philips’ customer service, and were especially frustrated that they were provided with US-specific information that was not relevant to the Canadian context. Overall, these customer service interactions were time-consuming and energy-depleting.

> It was terrible. We were basically told there’s a problem. [Philips] could not tell us how common it was. They could not tell us what the expected health implications of it were. Or how likely our patients were to experience those things. We were not told what mitigation strategies they were working on, or how quickly they expected to be able to replace parts or come up with a solution. So, we were very much in the dark. I think it was not well-handled. (Interview 44, Sleep Physician)

> I think the reason why they were tight-lipped, is because with any equipment-related recalls like this, there’s going to be a class-action lawsuit. It’s inevitable. And so, as a result I think they – whatever they communicated – they’ve touched with their legal team first and the legal team is guiding this conversation. (Interview 46, Health System Leader)

Participants emphasized that confusion and uncertainty persisted as the recall went on, related to unclear communications that lacked essential information relevant to patient care and clinical decision-making. Vendors and sleep clinics were patients’ main source of information, but it was challenging to provide advice on next steps due to infrequent updates.

> […] the constant changing of the information that they [Philips] had as well was frustrating for everybody involved […] That was probably among the worst part is just the constant changing the story, “OK well it’s this, nope, now we’re going to do this. Nope, now we’re going to do this.” (Interview 4, Vendor RT (Clinical))

#### Alternative sources of information to fill gaps

Participants from all groups reported difficulty in obtaining accurate and timely information. Several Canadian healthcare bodies responded by producing a joint position statement and guidance to help inform clinical decisions.

> I appreciated the guidance given by […] the Canadian Thoracic Society and the Canadian Sleep Society and, you know, the information that was provided to sleep physicians as guidance to how to handle the recall and, you know, what to do. (Interview 5, Sleep Physician)

Healthcare providers expressed a desire for greater communication from regulators such as Health Canada, the ability to engage with regulators to ask questions or express concerns, and for Health Canada to compel better communication from manufacturers during device recalls.

### Domain 2: Recall Execution

#### Notification and Traceability

Users of PAP therapy were typically notified of the recall by the vendor who had sold them their device, via written or electronic communication.

> We immediately pulled huge mailing lists, went through them, pulled all the recalled serial numbers out of our systems, and did mailers to our patients. That was the fastest way. And then they would have a written communication, too. So we did that for anyone who had purchased – all the way back to 2009 – equipment. So that was out and gone by the first week of July. (Interview 17, Vendor RT (Clinical))

While many individuals with OSA acknowledged the great lengths taken by vendors to notify their patients and help them understand their options, some perceived vendors to have performed the bare minimum or have downplayed the seriousness of the recall.

> Nowhere in this form letter does it identify which machines. […] You’re sending me a direct letter about this, but you are not explaining the basis for the recall. You’re telling me through the wording, I’m getting the impression that [my vendor’s] not really supporting this recall notice. “Hey, [patient name], there’s a recall notice.” Is it even my machine? It doesn’t say anything. (Interview 21, Individual with OSA)

Specialist sleep centres also notified their patients of the recall, and often directed patients to their websites for information and guidance. individuals with OSA reported booking specialist appointments specifically to discuss the recall. Some individuals with OSA indicated that they were never notified of the recall by their vendors or healthcare providers.

#### Device Replacement

Participants from all groups described the device replacement process as protracted, confusing, and constantly changing. It was unclear why replacement units were provided through a vendor rather than directly from the manufacturer, and lack of clarity around which model they would receive left many uncertain and anxious. Others were confused about why some devices instead had internal parts replaced and questioned the efficacy of this more limited remediation. Individuals with OSA were frustrated by extremely long wait times for replacement; at the time of interviews, some had been waiting over two years.

> It’s been extremely frustrating. And for myself, luckily, I still had my first machine that’s well over ten years old, […] so I was able to carry on without any issues. […] But the fact that I’m still out of pocket for that machine that I just replaced because they [Philips] never stepped up and delivered my machine. (Interview 26, Individual with OSA)

Vendors also described numerous frustrations, including: unclear and changing timelines; confusion over how replacements were prioritized; and logistical difficulties such as commercial deliveries arriving at retail locations and the burden of inventory management. Staff also devoted substantial amounts of time contacting patients, setting up new devices, and educating patients on new device models that were unfamiliar to them.

> So, some of the machines they’re [Philips] replacing the whole unit, some machines they’re only replacing the actual blower part, like the CPAP itself. And there’s no rhyme or reason. It doesn’t matter if you bought your machine two years ago or five years ago. Like there’s no rhyme or reason. (Interview 23, Vendor RT (Administrative))

> It’s a lot of different features on the machines, too, that we have to set it up. So, it’s a bit more complicated. And if they exchange for a completely different device, we also have to show them how to use it. We cannot just say, “There is the machine, good luck.” (Interview 10, Vendor RT (Clinical))

### Domain 3: Experience of the Recall

#### Impact on Individuals with OSA

The reasons for the recall caused deep concern among individuals with OSA about the potential adverse health risks (especially cancer). Those with recalled devices described a difficult choice between discontinuing or reducing therapy until device replacement or continuing to use their recalled device (against the manufacturer’s initial recommendation).

Even after making a choice, individuals who stopped or reduced therapy described anxiety around the implications of untreated OSA and many reported adverse effects on sleep quality, daytime function, and quality of life. Those who continued therapy described anxiety around increased exposure to a potentially harmful therapy. The uncertainty around the actual risk of negative health effects heightened feelings of stress.

> It made me very anxious trying to decide whether to use it or not […] normally I would just go to sleep and it was kind of a normal thing, but after finding this out about the particles, you know, potentially, you’d lay there and worry about it, you know, “is it shooting plastic particles into my lungs?” And then you either fall asleep or take it off, depending on how exhausted I was. (Interview 47, Individual with OSA)

A small minority of individuals with OSA expressed minimal concern, viewing the recall as a precautionary measure or reserving judgment until better evidence became available.

Individuals with OSA incurred both monetary and non-monetary costs due to the recall. Some paid for a new device rather than wait for a replacement, but this option was not financially feasible for many. Individuals living in rural and remote areas, where access to sleep care was limited, often drove long distances and sometimes stayed overnight in a hotel to visit a CPAP vendor or consult with a sleep specialist. Non-monetary costs included time demands such as arranging appointments and referrals with multiple providers.

> I’ve probably spent easily 20 hours talking to people on the phone and emails and stuff like that, to try to get somewhat of a resolution, and I’m no further ahead. (Interview 26, Individual with OSA)

#### Challenges for Healthcare Providers and Vendors

Healthcare providers reported lengthy conversations with patients about the choice to continue or stop therapy. They expressed concern for their patients’ wellbeing, especially those with severe disease who had chosen to stop therapy or could not access a non-recalled device. Healthcare providers and vendors experienced moral distress related to providing a potentially harmful treatment and being unable to offer an alternative. This experience led many to consider leaving their profession.

> I directly sell equipment to patients and just knowing that I sold them a device that is potentially harmful to them. You know they trusted me with their care, they trusted me to sometimes help them make that decision about equipment they’re purchasing long-term, and to hear that now all that equipment is being recalled and I sold it in good faith to these people, you know that didn’t feel great. (Interview 30, Vendor RT (Clinical))

Vendors felt the recall process shifted much of the responsibility for device replacements onto their businesses, which increased workload and financial burden. The requirement for vendors to facilitate device replacement also placed them in a difficult position between patients, providers, and the manufacturer.

> I had nothing to offer the patients. I had no solutions. As therapists, RTs, we’re critical thinkers, we’re problem solvers. And I had no solution. I have thousands of patients that I was looking to help, and I couldn’t. So it really was disheartening. (Interview 12, Public Sector RT (Clinical))

Vendor staff reported feeling “caught in the middle” and becoming targets for the anger and frustrations of all sides. Some vendors noted that Philips benefited from this arrangement, as it shielded them from direct patient interactions and because many patients assumed the recall was the vendors’ fault since vendors were the primary point of contact.

> We’ve taken a lot of abuse from people, verbal abuse from people that have come in. And it’s unfortunate, because I don’t have any answers. But I would say the most that we’ve gotten from them is just being so angry. (Interview 15, Vendor RT (Clinical))

### Domain 4: Policy and Regulatory Context

#### Trust

The recall and its execution damaged trust in both Philips and Canadian regulators. Many individuals with OSA switched or planned to switch to competitors. Healthcare providers and vendors reported reluctance to stock or recommend Philips devices. Trust in medical device regulations in general and Health Canada in particular was low, with some noting the device issues were not identified in Canada and criticizing Health Canada for largely following FDA decisions. The participants’ limited trust in Health Canada was largely fatalistic: they felt forced to rely on the system blindly due to a lack of transparency and no opportunity to independently assess regulatory decisions.

> I felt that the information a person was going to be given was the information they were going to be given, and you either had to trust it in blind faith or distrust it, just for the sake of – you know, just because. I didn’t feel that I had a whole lot of options […] (Interview 24, Individual with OSA)

Many participants emphasized that their loss of trust was not due to device problems alone but to a poor and disorganized response to remedying those problems. In contrast, a few participants felt the recall indicated the system was working: a problem was detected and steps taken to remediate it.

#### Transparency & Regulatory Effectiveness

Participants felt the recall lacked transparency from both Philips and Health Canada. This assessment was compounded by poor communication from both parties, leading to confusion about the recall’s execution and rationale for related decisions.

> Honestly, you put this thing on your face every night for eight hours, breathing it in. […] I think it would have been full disclosure. And you know what, that’s what I would say: it always felt like they [Philips] were trying to – I don’t know if it felt like they were trying to hide something, but they just weren’t being fully transparent, that’s what it felt like. (Interview 24, Individual with OSA)

Participants were particularly frustrated with what they perceived as Health Canada’s persistent silence and inaction following the initial recall announcement. Health Canada representatives, in written statements to the researchers, described how their actions met the requirements of applicable legislation and noted that they understood Philips’ actions to have done the same: “Phillips [sic] has provided all the information as required in [Canada’s] Medical Devices Regulations” (Health Canada, see Table 5 for full quote). However, participants stressed that “the regulations themselves could be improved” (Interview 14, Health System Leader), particularly around enforcement and the ability to compel manufacturer behaviour. The general sentiment was that current regulations were inadequate to serve the interests of device users and may have been counterproductive: multiple participants highlighted that the confusion experienced by device users and strain on vendors was a direct result of current regulations and “the Canadian government” regulating devices such that replacements effectively “had to go through a distributor” (Interview 23, Vendor (Administrative)).

## DISCUSSION

In this qualitative study, we explored the experiences of individuals from several groups affected by or involved with the Philips Respironics PAP recall, including those with OSA, healthcare providers, PAP vendors and health system leaders. Participants uniformly described significant problems with communication about the recall, initially and throughout the device replacement process. The identification of individuals with recalled devices and execution of mitigation processes were also confusing and frustrating. Importantly, the recall placed an exceptional burden on all stakeholders, with meaningful adverse effects on patient health and wellness, healthcare provider morale, and strain on vendor operations and financial viability. This is the first study to directly explore the experiences and perspectives of those affected by this large medical device recall.

Our results support the findings of two recent survey studies exploring the recall’s impacts on individuals with OSA and their healthcare providers. Robbins *et al*. surveyed 136 sleep clinicians who were American Academy of Sleep Medicine members.[25] Participants reported several ways that clinicians and patients learned about the recall; notably, only 7.9% of patients learned about the recall directly from Philips Respironics, whereas 45% became aware via social or news media. Most clinicians recommended that patients continue using their recalled device after consideration of relevant clinical factors, and approximately 60% of patients continued use. More recently, we surveyed 632 Canadians with OSA, and determined that over 15% of respondents were unaware of the recall, even two years after it began. Similarly, nearly half of survey participants discontinued or reduced PAP use.[26] In both studies, the recall and delays for device mitigation adversely impacted the health and well-being of both clinicians and patients, and eroded trust in medicine and the healthcare team.[25,26] In the present study, we identified similar themes related to poor communication, challenges with navigating the recall, and the resulting strain on clinicians. Our study extends the findings of prior survey studies by characterizing the burden of the recall on several groups, including individuals with OSA who used PAP, in greater depth.

Our study and others have highlighted several potential strategies to mitigate the impact of future recalls. First, device traceability and patient notification were key problem areas. Philips Respironics relied on PAP vendors and clinicians to notify patients, citing the inability to track PAP devices.[21,30] However, inadequate information to guide frontline providers led to confusion, delays, and situations where PAP users were either unaware of the recall or unable to make informed decisions about ongoing PAP use. Some clinics took steps to mitigate these challenges such as standardizing communication and developing decision support resources for staff and patients.[31] Moreover, medical societies and patient advocacy groups established guidance documents,[22,32–35] which improved the information environment but did not eliminate the strain experienced by providers nor the significant distress reported by affected individuals with OSA. Our findings support the adoption of reforms to address these problems, such as a central PAP device registry or modification of privacy legislation to enable tracing using unique device identifiers (UDIs), similar to other medical devices.[36] Once recalled devices have been identified, studies of implantable cardiac device recalls have identified the need for ongoing patient support and clear communication.[37,38] This could be a collaborative effort between device manufacturers, vendors, healthcare providers and medical or patient societies.

Unlike implanted devices, PAP devices may be owned by multiple individuals over time. The mostly private funding of OSA treatment in several Canadian provinces,[27] combined with a vibrant PAP resale market, presents a major barrier to traceability of recalled devices and notification of affected individuals. With increasing internet connectivity of current PAP devices, manufacturers could address this issue by incorporating digital recall notifications into device interfaces. Moreover, regulatory bodies could mandate this function as a condition of authorization. A prior field safety notice of ResMed’s Adaptive Servo-Ventilation devices for individuals with central sleep apnea and left ventricular dysfunction presented a similar risk of severe adverse outcomes.[39] In that case, however, the number of affected individuals was much lower and most were followed in specialized sleep or cardiology clinics. The scale of the Philips recall is an order of magnitude larger, thus necessitating different solutions for traceability and notification.

Second, participants described difficulties in obtaining replacement devices due to an opaque device registration process, uncertainty about the nature of recall mitigation (e.g., repair vs replacement) and inaccurate timelines. Logistical challenges related to device inventory placed an avoidable burden on vendors, which was exacerbated by difficulty obtaining devices from other manufacturers due to supply chain issues. A consistent message from our study and others was that a more concerted effort is required to organize timely, safe and comprehensive execution of a medical device recall.[21,25,30,40] Potential solutions include government-mandated processes, timelines for recall mitigation (tied to regulatory device approvals) and restrictions on manufacturer-imposed deadlines for registering recalled devices (device registration in Canada and the United States ended on December 31, 2024, but Philips may honour remediation requests if contacted directly [personal communications with Health Canada and Philips]).[41,42] Such regulatory changes must be balanced to avoid discouraging PAP manufacturers from marketing products in Canada. [43]

Third, the policy environment for OSA care hindered the timeliness and effectiveness of recall execution. There is wide variation in OSA care delivery, funding and regulations across Canadian provinces and territories.[27] Thus, while the recall was announced by a federal agency (Health Canada), and despite existing medical device regulations and licencing requirements, implementation either could not be standardized or was not enforced, which we propose contributed to confusion around accountability for various aspects of the recall. If an individual has moved between provinces or their healthcare providers or vendor are no longer available, notification about and navigation of the recall would be extremely difficult. A detailed discussion of these issues is beyond the scope of this paper, but strategies such as nationally standardized recall procedures, federal negotiation or oversight of device manufacturer contracts to require more direct engagement in recall activities, and consistent public funding of PAP devices to enable enforcement of regulations could mitigate these system barriers.[27]

This study’s strengths include consideration of multiple perspectives representing most Canadian provinces and territories. We also tailored interviews to each participant group while maintaining common themes to strengthen the analysis. Finally, our approach enabled a deep exploration of participant perspectives to characterize the recall’s wide-ranging effects. This study also has limitations which include potential volunteer bias: individuals with greater challenges navigating the process or distress from the recall may have been more likely to participate, leading to overestimation of the recall’s adverse impacts. The timing of interviews (17-26 months after the recall announcement) raises the possibility of recall bias and does not capture the participants’ dynamic experiences as the recall unfolded. Individual experiences with the recall may be influenced by local factors such as the model of OSA care or funding for PAP therapy,[27,28] raising a threat to generalizability that could have been exacerbated by the relatively small sample in some groups (e.g., physicians, health system leaders). However, we found perspectives to be consistent across provinces, and our results also mirror those of studies in other countries.[21,25,31,32] Finally, we did not interview representatives from Philips Respironics to obtain their perspectives on the recall.

In summary, the Philips Respironics recall had serious consequences for affected individuals and organizations, related to communication, execution, and policy. These findings underscore the need to improve strategies for managing large-scale medical device recalls.

## Supporting information

SRQR Checklist

## CONTRIBUTIONS (CRediT statement)

**Sachin R. Pendharkar:** Conceptualization, Funding acquisition, Supervision, Methodology, Writing - Original Draft, Writing - Review & Editing

**Kenneth G. Blades:** Methodology, Project administration, Investigation, Data Curation, Formal analysis, Software, Writing - Original Draft, Writing - Review & Editing

**Bashour Yazji:** Formal analysis, Writing - Original Draft, Writing - Review & Editing

**Najib T. Ayas:** Funding acquisition, Resources, Writing - Review & Editing

**Robert L. Owens:** Resources, Writing - Review & Editing

**Marta Kaminska:** Resources, Writing - Review & Editing

**Constance Mackenzie:** Resources, Writing - Review & Editing

**Andrea S. Gershon:** Resources, Writing - Review & Editing

**Diana C. Ratycz:** Methodology, Writing - Review & Editing

**Vadym Lischenko:** Methodology, Writing - Review & Editing

**Mark E. Fenton:** Resources, Writing - Review & Editing

**Kerry A. McBrien:** Methodology, Resources, Writing - Review & Editing

**Marcus Povitz:** Resources, Writing - Review & Editing

**Tetyana Kendzerska:** Conceptualization, Funding acquisition, Supervision, Methodology, Writing - Original Draft, Writing - Review & Editing

## STATEMENTS AND DECLARATIONS

### Research Ethics

Institutional review board approval was obtained from the University of Calgary Conjoint Health Research Ethics Board (REB22-0803), the Ottawa Hospital Research Institute (Ethics ID: 20220590-01T), and Clinical Trials Ontario (Ethics ID: CTO-4064).

### Informed Consent

All participants provided informed consent to participate via a signed consent form and/or explicit verbal consent.

### Data Availability

The data underlying this study cannot be shared due to research ethics board requirements which protect the confidentiality of the research participants.

### Reporting Guidelines

Study design was informed by the Standards for Reporting Qualitative Research (SRQR) and the Consolidated Criteria for Reporting Qualitative Research (COREQ). An SRQR checklist is included alongside the manuscript.

### Author Approvals

All authors have seen and approved this manuscript.

