## Supplement for "Exploring the Impact of a Medical Device Recall on Individuals with Obstructive Sleep Apnea and Healthcare Providers: A Qualitative Study"

### ONLINE DATA SUPPLEMENT

#### 1. Summary of organizations and channels engaged for participant recruitment

**Medical Organizations**

- Canadian Sleep Society
- Canadian Thoracic Society
- The Lung Association (all provincial/territorial chapters)
- Fondation Sommeil
- Canadian Society of Respiratory Therapists
- Alberta Health Services Strategic Clinical Network Patient and Family Engagement Office
- Alberta Medical Association
- Alberta College of Family Physicians
- Association pulmonaire du Quebec

**Public Programmes**

- Ontario Ventilator Equipment Pool
- Alberta Respiratory Benefits Program
- British Columbia Provincial Respiratory Outreach Program
- Quebec National Program for Home Ventilatory Assistance
- Quebec Committee for Home Ventilation
- Saskatchewan Aides to Independent Living Program

**Academic Venues**

- Strategy for Patient Oriented Research Support Units (Ontario, Alberta)
- Departments of Family Medicine (Ottawa, Calgary)
- Canadian Sleep and Circadian Network
- Canadian Sleep Research Consortium

**Clinics, Equipment Vendors**

- PAP vendors, suppliers, respiratory homecare companies
- Public sleep laboratories
- Private sleep laboratories

**Social Media / Online Platforms**

- All organizations above if active on social media
- Study investigators' social media accounts
- Sleep On It (sleeponitcanada.ca)

**2. Interview Guide for Individuals with OSA**

**Participant Background**
Relevant information and characteristics about the participant.

1. **Have you been prescribed a form of PAP therapy? (i.e., “breathing machine”)**
    a. What type?
        - CPAP, BPAP, APAP, other, multiple
    b. When?
        - *Suggested probes:*
        - Date of initial prescription
        - Date of treatment start (before/after June 2021)
        - Roughly how long have you been using PAP?
    c. Are you currently using PAP therapy?
        - *If yes:* About how often?
        - *If no:* When did you stop? When you did use PAP, about how often?
        - *Suggested probes re frequency:*
        - Hours per night
        - Nights per week
    d. [ *Definitions, if participants need clarification*: ]
        - PAP = positive airway pressure (“breathing machine”)
        - CPAP = continuous
        - BPAP = bi-level
        - APAP = auto-titrating, adjustable
2. **What do you use PAP therapy for?**
    a. *Alt phrasing:* What is your diagnosis?
        - *e.g.,* sleep apnea (OSA, CSA), COPD, heart/lung failure
    b. When, approximately, were you first diagnosed?
    c. How does your healthcare provider classify or describe your condition?
        - Mild, moderate, severe, other?
        - Do you also use oxygen?
    d. [ *Definitions, if participants need clarification*: ]
        - OSA: “obstructive,” airway blockages during sleep
        - CSA: “central,” signals from brain to lungs interrupted
        - COPD: various conditions where airflow blocked to lungs, not just during sleep
        - Resp Failure: PAP ventilation used when oxygen via facemask insufficient or impossible
3. **Can you tell me about your PAP device?**
    a. How did you get your current device?
        - Public insurance, private insurance, paid out of pocket, combination
        - Who wrote your PAP prescription? (e.g., sleep physician, other physician)

    - Do you have more than one device? (e.g., for travel, backup, cottage)
    - Have you ever replaced your device since you first started using one?
b. When did you acquire your current device?
    - *Alt phrasing:* How old is your current device?
c. Who is the manufacturer of your current PAP device?
    - Philips Respironics, ResMed, DeVilbiss, Fisher & Paykel, other
    - Is your device affected by the recall?

**I. Experience of Recall**

How the participant experienced and reacted to the recall.

**4. How did you first learn about the recall affecting Philips Respironics PAP devices?**

a. From whom did you first hear about the recall? How?
    - *e.g.,* Philips Respironics, PAP vendor, sleep physician, family physician, respiratory therapist, other patient(s), the news/media, social media

b. When did you first hear about the recall?
    - Year, month, day (if possible)

c. What information did you learn at this time?
    - Reason for recall
    - Scope of recall
    - Actions being taken as a result of the recall

d. Was it clear, upon first learning of the recall, whether or not you were directly affected by the recall?

**5. What was your reaction to the recall?**

e. What were your main questions or concerns?

f. Were you able to get answers to your questions or concerns?

**6. What else have you learned about the recall in the time since then?**

g. At what points did you learn new information about the recall?

h. What new information did you receive at these times?

i. From whom?

**7. Was the information you received about the recall useful?**

j. Did it provide you with sufficient knowledge and understanding of what was happening?

k. Did it address your questions and concerns?

**8. Was the communication around this recall useful?**

l. What communications did you find useful or helpful? Why?

m. What communications were <u>not</u> useful or made things worse? Why?

n. What could have made these communications better?
    - *Suggested Probes:*
    - Type of information supplied
    - Clarity, tone
    - Frequency, updates

- Recommended courses of action

o. [ *Note: This question may include communications from and with:* ]
   - Philips Respironics
   - PAP vendors, homecare companies
   - Healthcare providers (sleep physicians, family physicians, RTs)
   - Regulators, policymakers
   - News media

**II. Impact of Recall**

What effects the recall had upon the participant.

**9. How has this recall affected you?**

- *Suggested Probes:*

a. *Emotionally,* or in terms of your mental health

b. *Health,* such as:
   - stability or severity of your breathing disturbances during sleep
   - emergence of new symptoms, possible side effects of modifying or stopping PAP use (e.g., increased AHI, arrhythmia)
   - symptoms related to recall (e.g., respiratory irritation)

c. *Relationships* with healthcare providers (your doctor, vendor, RT)

d. *Confidence* in your prescribed therapy or healthcare system in general

**10. What, if any, decisions or actions did you need to take as a result of this recall?**

a. Consults with your usual care team? (family doctor, sleep doctor, RT)

b. Second opinions from professionals other than your usual care team?
   - *If yes:* Why did you seek a second opinion?
     [ *Intent:* was it patient driven or care-team-driven? ]

c. Did you make any changes to your usual PAP therapy?
   - *Examples of possible changes:*
     - Did you use you PAP device less frequently?
     - Did stop using your PAP device?

   i. *If no:* Did you consider doing so, or consult with a healthcare provider about doing so?

   ii. *If yes:* Did you do so in consultation with a healthcare provider?

   iii. *If yes:* How did these changes affect you?
     - *e.g.,* How long were you off your device?
     - *e.g.*, Did symptoms worsen as a result?
     - *e.g.,* Did this impact your work or quality of life?

**11. Do you think you had the information you needed to make the best decisions or take the best courses of action?**

- *If yes:* what information helped you do this and how?
- *If no:* what information would have helped you do this?

**12. Were there actions you wanted to take but could not?**

a. *e.g.,* replace PAP device but could not due to device shortage

**13. Did you incur any costs or expenses as a result of the recall?**

a. *e.g.,* repair or replace PAP device, out of pocket screening for possible negative health effects, etc.

**III. Suggestions for Improvement**

What the participant thinks could be done better next time, or in a similar situation.

**14. What could have been done differently to improve your experience with this recall?**

a. How could the communications have been better?
    - Clearer, faster, more/less frequent, more/less substantive, etc.

b. Who could have communicated better, and why/how?
    - Philips Respironics, healthcare providers, PAP vendors, regulators

c. What actions could or should have been taken, but were not?
    - By whom? (Philips Respironics, healthcare providers, vendors, regulators)

**15. If something like this were to happen again, what should be done about it?**

d. What should <u>manufacturers</u> of <u>affected devices</u> do?
    - *e.g.,* repair or replace devices, offer preventive screening, offer reimbursement or partial reimbursement schemes for out of pocket expenses, etc.

e. What should <u>manufacturers</u> of <u>non-affected devices</u> do?
    - *e.g.,* notify users to re-assure they are not affected, subsidise cost of their own devices for users of affected devices seeking replacement, etc.

f. What should <u>healthcare providers</u> do?
    - Individual healthcare professionals and clinics (e.g., your family doc or local clinic)
    - Healthcare associations and colleges (e.g., Canadian Medical Assoc, if pt asks for eg)

g. What should <u>PAP vendors</u> and <u>homecare companies</u> do?
    - *e.g.,* establish/maintain lines of communication between patients, manufacturers, and healthcare providers/associations
    - *e.g.,* help repair or source replacement devices for affected users

h. What should provincial or national <u>regulators</u> and <u>policymakers</u> do to help prevent recalls like this, or manage them better when they happen?
    - *e.g.,* establish/maintain a post-market surveillance system for PAP devices
    - *e.g.,* set manufacturer standards regarding communication, repair, replacement, compensation, etc., in event of recall, and audit compliance

**16. When it comes to telling patients about a medical device recall, who do you think should be primarily responsible to do this?**

i. Who should the messenger be?
    - *e.g.,* manufacturer, healthcare professionals, a combination?

j. What is the best way to do so?
    - Content, framing, method of delivery, etc.

**17. Overall, how would you improve your experience with this recall?**

Thank you for taking the time to help us with this work. Before we conclude: is there anything you would like to add — or any suggestions or recommendations you would like to make — that we have not asked you about?

**3. Interview Guide for Healthcare Providers (Physicians and Respiratory Therapists)**

**Participant Background**
Relevant information and characteristics about the participant.

1. **What is your role?**
    a. Physician, respiratory therapist, other?
    b. How many years have you been in practice?
    c. Do you have any sleep-specific training? (e.g., sleep specialization, sleep-related CME, etc.)

2. **Where do you currently practice?**
    a. *Location:* urban, rural, remote
    b. *Type:* public or private, setting (clinic, hospital, sleep centre)

3. **Can you tell us a bit about the nature of your practice and patient population?**
    a. Approximately how many patients do you have?
        - On average, about how many patients do you see per week?
        - About how many of your patients have an OSA diagnosis?
        - About how many of your patients have OSA suspected or probable?
    b. How would you characterize your overall patient population?
        - *In terms of:* demographics, SES, social determinants of health, etc.
    c. How would you characterize your OSA patients?
        - *In terms of:* severity, complexity, comorbidities, etc.

**I. Experience of Recall**
How the participant experienced and reacted to the recall.

4. **How did you first learn about the recall affecting Philips Respironics PAP devices?**
    a. From whom did you first hear about the recall? How?
        - *e.g.,* Philips Respironics, PAP vendor, sleep physician, family physician, respiratory therapist, other patient(s), the news/media, social media
    b. When did you first hear about the recall?
        - Year, month, day (if possible)
    c. What information did you learn at this time?
        - Reason for recall
        - Scope of recall
        - Actions being taken as a result of the recall
    d. Was it clear, upon first learning of the recall, whether or not any of your patients were directly affected by the recall?
5. **What was your reaction to the recall?**
    e. What were your main questions or concerns?
    f. Did you take any actions as a result of learning about the recall? (e.g., contact colleagues, healthcare associations, PAP vendors, etc.)

6. **Did any of your patients contact you about the recall?**
   g. *If yes:*
      - How would you say the recall notice affected your patients?
      - What were your patients' primary concerns?
      - Did any of your patients discuss modifying or ceasing PAP therapy?
   h. *If no:*
      - Did you anticipate or prepare for such conversations with patients?
      - What topics did you anticipate, and how did you prepare?
   i. In general, what recommendations did you make to your patients who were affected by, or otherwise concerned about, the recall?
   j. Can you describe how you arrived at these recommendations?
   k. Did you initiate contact with any of your patients about the recall?
      - How? [ *Intent:* feasibility of IDing pts, time required to contact, etc. ]
      - Why? [ *Intent:* intrinsic motivation, Health Canada responsibility, etc. ]
7. **How would you assess the information you have received about the recall?**
   l. Did it provide you with sufficient knowledge and understanding of what was happening?
   m. Did it address your questions and concerns?
   n. Did it address the questions and concerns of your patients?
   o. Did it provide you with sufficient means to communicate effectively with your patients?
8. **How would you assess the quality of communication around this recall?**
   p. What communications did you find useful or helpful? Why?
   q. What communications were <u>not</u> useful or made things worse? Why?
   r. What could have made these communications better?
      - *Suggested Probes:*
      - Type of information supplied
      - Clarity, tone
      - Frequency, updates
      - Recommended courses of action
   s. [ *Note: This question may include communications from and with:* ]
      Philips Respironics; PAP vendors, homecare companies; Healthcare providers (sleep physicians, family physicians, RTs); Regulators, policymakers; News media

**II. Impact of Recall**
What effects the recall had upon the participant.

9. **How has this recall affected you?**
   - *Suggested Probes:*
   a. *Professionally,* in terms of your ability to serve your patients
   b. *Relationships* with your patients
      - *e.g.,* your patients' trust/confidence in:
        1. Device manufacturers
        2. PAP therapy

    3. The healthcare system
    4. You
c. *Relationships* with other healthcare providers (e.g., your clinic staff, colleagues, etc.)
d. *Relationships* with external service providers (e.g., referral centres such as sleep centres, PAP vendors, etc.)
e. *Relationships* with policymakers (e.g., levels of engagement, trust, communication)
f. *Workload:* how much extra time did you spend on this issue?
    - *e.g.,* contacting patients, colleagues, vendors, regulators, etc.

**10. What, if any, decisions or actions did you need to take as a result of this recall?**

a. Consultations or meetings with colleagues at your clinic or institution, or other clinics or institutions?
b. Requests for information from device manufacturers, healthcare associations or colleges, regulators/policymakers, etc.?
c. Did you consider, independently or in consultation with fellow professionals, any of these courses of action:
    - Modifying the frequency or duration of device usage for your PAP patients, or a subset of these patients?
    - Treatment cessation (including temporary cessation) for your PAP patients, or a subset of these patients?
    - What factors did you weigh when considering these or similar courses of action?
    - What factors did you weigh when making recommendations to patients who asked, or may have asked, about such courses of action?
d. Do you think you had the information you needed to make the best decisions or take the best courses of action?
    iv. *If yes:* what information helped you do this and how?
    v. *If no:* what information would have helped you do this?

**III. Suggestions for Improvement**

What the participant thinks could be done better next time, or in a similar situation.

**11. What could have been done differently to improve your experience with this recall?**

a. How could the communications have been better?
    - Clearer, faster, more/less frequent, more/less substantive, etc.
b. Who could have communicated better, and why/how?
    - Philips Respironics, healthcare providers, PAP vendors, regulators
c. What actions could or should have been taken, but were not?
    - By whom? (Philips Respironics, healthcare providers, vendors, regulators)

**12. If something like this were to happen again, what should be done about it?**

d. What should <u>manufacturers</u> of <u>affected devices</u> do?

- *e.g.,* repair or replace devices, offer preventive screening, offer reimbursement or partial reimbursement schemes for out of pocket expenses, etc.

e. What should <u>manufacturers</u> of <u>non-affected devices</u> do?
   - *e.g.,* notify users to re-assure they are not affected, subsidise cost of their own devices for users of affected devices seeking replacement, etc.

f. What should <u>healthcare providers</u> do?
   - Individual healthcare professionals and clinics (e.g., family physicians or local clinics)
   - Healthcare associations and colleges (e.g., CSCN, CMA, etc.)

g. What should <u>PAP vendors</u> and <u>homecare companies</u> do?
   - *e.g.,* establish/maintain lines of communication between patients, manufacturers, and healthcare providers/associations
   - *e.g.,* help repair or source replacement devices for affected users

h. What should provincial or national <u>regulators</u> and <u>policymakers</u> do to help prevent recalls like this, or manage them better when they happen?
   - *e.g.,* establish/maintain a post-market surveillance system for PAP devices
   - *e.g.,* set manufacturer standards regarding communication, repair, replacement, compensation, etc., in event of recall, and audit compliance

**13. When it comes to telling patients about a medical device recall, who do you think should be primarily responsible to do this?**

i. Who should the messenger be?
   - *e.g.,* manufacturer, healthcare professionals, a combination?

j. What is the best way to do so?
   - Content, framing, method of delivery, etc.

**14. Overall, how would you improve your experience with this recall?**

Thank you for taking the time to help us with this work. Before we conclude: is there anything you would like to add — or any suggestions or recommendations you would like to make — that we have not asked you about?

### 4. Interview Guide for Health System Leaders

#### Participant Background

Relevant information and characteristics about the participant.

1. **What is your role?**
    a. Policy, regulation, other?
    b. In what capacity?
        - Government, industry, insurance, other?
        - *Scope:* national, provincial, other?
    c. How many years have you been in your role?

2. **What is your relationship to the world of sleep medicine and PAP therapy?**
    a. What contacts and interactions do you have with industry?
        - *e.g.,* device manufacturers, vendors, insurance providers, etc.
    b. What contacts and interactions do you have with healthcare professionals?
        - *e.g.,* sleep specialists, respirologists, primary care providers, RTs
    c. What contacts and interactions do you have with sleep apnea patients?
        - *Specifically:* OSA patients undergoing PAP therapy

3. **What is your relationship to the recall of Philips Respironics PAP devices?**
    a. Were you involved in issuing the recall notice?
    b. Were you involved in disseminating the recall notice?
    c. Were you involved in interpreting the recall notice?
        - *e.g.,* to brief government decisionmakers, healthcare associations, patient representation groups, or other stakeholder audiences?
    d. Were you involved in planning a response to the recall?

#### I. Experience of Recall

How the participant experienced and reacted to the recall.

4. **How did you first learn about the recall affecting Philips Respironics PAP devices?**
    a. From whom did you first hear about the recall? How?
        - *e.g.,* Philips Respironics, PAP vendor, sleep physician, family physician, respiratory therapist, other patient(s), the news/media, social media
    b. When did you first hear about the recall?
        - Year, month, day (if possible)
    c. What information did you learn at this time?
        - Reason for recall
        - Scope of recall
        - Actions being taken as a result of the recall
    d. Was it clear, upon first learning of the recall, whether or not any of your areas of responsibility were directly affected, or potentially affected, by the recall?
5. **What was your reaction to the recall?**

e. What were your main questions or concerns?
f. Did you take any actions as a result of learning about the recall? (e.g., contact colleagues, healthcare associations, PAP vendors, other regulatory bodies, industry contacts/reps, etc.)

6. **Did any of your stakeholders contact you about the recall?**
   g. *If yes:*
      - How would you say the recall notice affected your stakeholders?
      - What were your stakeholders' primary concerns?
      - Did any of your stakeholders discuss what advice or recommendations they should offer to their own stakeholders and/or patients?
   h. *If no:*
      - Did you anticipate or prepare for such conversations?
      - What topics did you anticipate, and how did you prepare?
   i. In general, what recommendations did you make to your stakeholders who were affected by, or otherwise concerned about, the recall?
   j. Can you describe how you arrived at these recommendations?
7. **How would you assess the information you have received about the recall?**
   k. Did it provide you with sufficient knowledge and understanding of what was happening?
   l. Did it address your questions and concerns?
   m. Did it address the questions and concerns of your stakeholders?
   n. Did it provide you with sufficient means to communicate effectively with your stakeholders or otherwise perform your role?
8. **How would you assess the quality of communication around this recall?**
   o. What communications did you find useful or helpful? Why?
   p. What communications were <u>not</u> useful or made things worse? Why?
   q. What could have made these communications better?
      - *Suggested Probes:*
      - Type of information supplied
      - Clarity, tone
      - Frequency, updates
      - Recommended courses of action
   r. [ *Note: This question may include communications from and with:* ]
      Philips Respironics; PAP vendors, homecare companies; Healthcare providers (sleep physicians, family physicians, RTs); Regulators, policymakers; News media

**II. Impact of Recall**
What effects the recall had upon the participant.

9. **How has this recall affected you?**
   - *Suggested Probes:*
   a. *Professionally,* in terms of your ability to serve your stakeholders
   b. *Relationships* with your primary stakeholders
      - *e.g.,* your stakeholders' trust/confidence in:

        1. Device manufacturers
        2. PAP therapy
        3. The healthcare system
        4. The medical device industry
        5. You
    c. *Relationships* with other regulators, policymakers, or decisionmakers
    d. *Relationships* with healthcare providers
    e. *Relationships* with patients
    f. *Workload:* how much extra time did you spend dealing with this issue?
        - e.g., meetings and consultations with providers, patients, officials, etc.

**10. What, if any, decisions or actions did you need to take as a result of this recall?**

a. Consultations or meetings with colleagues in your area of responsibility?
    - *e.g.,* government officials, ministry contacts, industry reps
b. Requests for information from device manufacturers, healthcare associations or colleges, regulators/policymakers, etc.?
c. Did you consider, independently or in consultation with fellow professionals, any of these courses of action:
    - Issuing public statements about the recall notice?
    - Issuing non-public, limited-circulation statements about the recall notice to relevant professional associations, institutions, or bodies?
    - Articulating an institutional position regarding the recall notice?
    - What factors did you weigh when considering these or similar courses of action?
    - What factors did you weigh when making recommendations to stakeholders who asked, or may have asked, about such courses of action, or who may have considered taking such action themselves?
d. Do you think you had the information you needed to make the best decisions or take the best courses of action?
    - vi. *If yes:* what information helped you do this and how?
    - vii. *If no:* what information would have helped you do this?

**III. Suggestions for Improvement**

What the participant thinks could be done better next time, or in a similar situation.

**11. What could have been done differently to improve your experience with this recall?**

a. How could the communications have been better?
    - Clearer, faster, more/less frequent, more/less substantive, etc.
b. Who could have communicated better, and why/how?
    - Philips Respironics, healthcare providers, PAP vendors, regulators
c. What actions could or should have been taken, but were not?
    - By whom? (Philips Respironics, healthcare providers, vendors, regulators)

**12. If something like this were to happen again, what should be done about it?**

d. What should <u>manufacturers</u> of <u>affected devices</u> do?

- *e.g.,* repair or replace devices, offer preventive screening, offer reimbursement or partial reimbursement schemes for out of pocket expenses, etc.

e. What should manufacturers of non-affected devices do?
   - *e.g.,* notify users to re-assure they are not affected, subsidise cost of their own devices for users of affected devices seeking replacement, etc.

f. What should healthcare providers do?
   - Individual healthcare professionals and clinics (e.g., family physicians or local clinics)
   - Healthcare associations and colleges (e.g., CSCN, CMA, etc.)

g. What should PAP vendors and homecare companies do?
   - *e.g.,* establish/maintain lines of communication between patients, manufacturers, and healthcare providers/associations
   - *e.g.,* help repair or source replacement devices for affected users

h. What should provincial or national regulators and policymakers do to help prevent recalls like this, or manage them better when they happen?
   - *e.g.,* establish/maintain a post-market surveillance system for PAP devices
   - *e.g.,* set manufacturer standards regarding communication, repair, replacement, compensation, etc., in event of recall, and audit compliance

**13. When it comes to telling patients about a medical device recall, who do you think should be primarily responsible to do this?**

i. Who should the messenger be?
   - *e.g.,* manufacturer, healthcare professionals, a combination?

j. What is the best way to do so?
   - Content, framing, method of delivery, etc.

**14. Overall, how would you improve your experience with this recall?**

Thank you for taking the time to help us with this work. Before we conclude: is there anything you would like to add — or any suggestions or recommendations you would like to make — that we have not asked you about?
