## Supplementary material for "Exploring the Impact of a Medical Device Recall on Individuals with Obstructive Sleep Apnea and Healthcare Providers: A Qualitative Study": SRQR Checklist

### Checklist — Standards for Reporting Qualitative Research (SRQR)

| Item | Description | Location (or reason for not reporting) |
|---|---|---|
| **Title & Abstract** | | |
| Title | Describe the nature and topic of the study. Identify the study as qualitative or indicate the approach or data collection methods. | Title, p. 1 |
| Abstract | Summarise the key elements of the study using the abstract format of the intended publication. | Abstract, p. 4 |
| **Introduction** | | |
| Problem Formulation | Describe the problem/phenomenon studied, its significance, relevant theory and empirical work, and gaps in current knowledge. | Abstract, p. 4<br>Introduction, pp. 5-6 |
| Purpose or research question | Describe the purpose of the study and specific objectives or questions. | Abstract, p. 4<br>Introduction, pp. 5-6 |
| **Methods** | | |
| Qualitative approach and research paradigm | Describe your qualitative approach, your guiding theory (if appropriate), and research paradigm, and reasons for your choices. | Study Design, pp. 6-7 |
| Researcher characteristics and reflexivity | Describe how researchers' characteristics may influence the research, including personal attributes, qualifications/experience, relationship with participants, assumptions, and/or presuppositions; potential or actual interaction between researchers' characteristics and the research questions, approach, methods, results and/or transferability. | Data Collection, p. 9<br>Data Analysis, pp. 9-10 |
| Context | Describe the setting/site(s) in which the study was conducted, why it was selected, and any other salient contextual factors that may influence the study. | Study Setting, p. 7<br>Study Participants, pp. 7-8 |
| Sampling strategy | Describe how and why research participants, documents, or events were selected; criteria for deciding when no further sampling was necessary, and the rationale for those criteria. | Study Participants, pp. 7-8<br>Data Collection, p. 9 |
| Ethical issues pertaining to human subjects | Describe any approval by an appropriate ethics review board and participant consent, or explain any lack thereof. Describe any other confidentiality and data security issues. | Declarations, p. 3<br>Study Design, pp. 6-7<br>Data Collection, p. 9 |
| Data collection methods | Describe the types of data collected; details of data collection procedures including (as appropriate) start and stop dates of data collection and analysis, iterative process, triangulation of sources/methods, and modification of procedures in response to evolving study findings. Describe your rationale for these choices. | Data Collection, p. 9<br>Online Supplement, pp. 1-14 |
| Data collection instruments and technologies | Describe any instruments (e.g., interview guides, questionnaires) and devices (e.g., audio recorders) used for data collection; describe if/how the instrument(s) changed over the course of the study. | Data Collection, p. 9<br>Online Supplement, pp. 1-14 |

| | | |
|---|---|---|
| Units of study | Describe the number and relevant characteristics of participants, documents, or events included in the study. Describe the level of participation. | Study Participants, pp. 7-8<br>Results, p. 10<br>Table 1, p. 26 |
| Data processing | Describe the methods for processing data prior to and during analysis, including transcription, data entry, data management and security, verification of data integrity, data coding, and anonymisation / deidentification of excerpts. | Data Collection, p. 9<br>Data Analysis, pp. 9-10 |
| Data analysis | Describe the process by which inferences, themes, etc. were identified and developed, including the researchers involved in data analysis; usually references a specific paradigm or approach. Describe why you chose this process. | Data Analysis, pp. 9-10 |
| Techniques to enhance trustworthiness | Describe any techniques to enhance trustworthiness and credibility of data analysis,(e.g., member checking, triangulation, audit trail). Describe why you chose these techniques. | Data Analysis, pp. 9-10 |
| **Results** | | |
| Synthesis and interpretation | Describe the main findings (e.g., interpretations, inferences, and themes); might include development of a theory or model, or integration with prior research or theory. | Results, pp. 10-17 |
| Links to empirical data | Provide evidence (e.g., quotes, field notes, text excerpts, photographs) to substantiate analytic findings. | Results, pp. 10-17<br>Tables 2-5, pp. 27-31 |
| **Discussion** | | |
| Integration with prior work, implications, transferability, and contribution(s) to the field | Summarize the main findings, explain how findings and conclusions connect to, support, elaborate on, or challenge conclusions of earlier scholarship; discuss the scope of application/generalizability; identify unique contribution(s) to scholarship in a discipline or field. | Discussion, pp. 17-21 |
| Limitations | Discuss the trustworthiness and limitations of findings | Discussion, p. 21 |
| **Other** | | |
| Conflicts of interest | Describe any potential sources of influence or perceived influence on study conduct and conclusions. Describe how these were managed. | Declarations, p. 3 |
| Funding | Describe sources of funding and other support. Describe the role of funders in data collection, interpretation, and reporting. | Declarations, p. 3 |

### Sources

O'Brien BC, Harris IB, Beckman TJ, Reed DA, Cook DA. Standards for reporting qualitative research: A synthesis of recommendations. Academic Medicine [Internet]. 2014 Sep;89(9):1245–51. Available from: https://journals.lww.com/academicmedicine/fulltext/2014/09000/Standards_for_Reporting_Qualitative_Research__A.21.aspx

O'Brien BC, Harris IB, Beckman TJ, Reed DA, Cook DA. The SRQR reporting checklist. In: Harwood J, Albury C, Beyer J de, Schlüssel M, Collins G, editors. The EQUATOR network reporting guideline platform [Internet]. The UK EQUATOR Centre; 2025. Available from: https:/resources.equator-network.org/guidelines/srqr/srqr-checklist.docx
